# Incremental costs of transitioning from four to eight WHO-recommended antenatal care visits in Uganda: A costing analysis from a societal perspective

**DOI:** 10.64898/2026.06.10.26355347

**Authors:** Elly B Atuhumuza, Esther C Atukunda, Angella Musiimenta, Godfrey R Mugyenyi, Jessica Haberer, Celestino Obua, Mark J Siedner, Lynn T. Matthews, Vincent Batwala, Van T Nghiem

## Abstract

**Background:** In 2016, the World Health Organization revised its antenatal care (ANC) recommendation from four to eight visits. For low- and middle-income countries like Uganda, where achieving even four visits remains a challenge, this transition has significant cost implications for both the health system and households. This study estimated the incremental costs of adopting the eight-visit model from a societal perspective.

**Methods:** The study was conducted in six government health facilities in southwestern Uganda. A micro-costing approach estimated health facility costs (personnel, equipment, consumables, and overhead). Costs incurred at patients’ end (transport, ultrasound, medical expenses, and time) were collected from 785 women using a questionnaire, with all costs in 2025 USD.

**Results:** For an average of 4.3 visits, total cost per woman was $100.1: facility costs $43.7 (43.7%), and patient costs $56.4 (56.3%). Transitioning to eight visits would increase total cost by $57.8 (57.8%), of which $36.4 (63.0%) would fall on households, equivalent to 68.8% of average monthly household income. Total costs would rise by 55.4% ($115.5 to $179.5) at Health Center IVs and 64.3% ($102.3 to $168.1) at Health Center IIIs, with facility costs up 43.4% and 62.9% and patient costs up 61.2% and 65.7%, respectively.

**Conclusion:** Transitioning to eight ANC visits would impose a large financial burden on households, with the incremental patient cost equivalent to more than two-thirds of average monthly household income. Equitable implementation requires improving availability of medicines and diagnostics, subsidizing transport, exploring telemedicine or community-based models, and improving efficiency at lower-tier health centers.

**Highlights:**

1. Evidence on the incremental costs of scaling up antenatal care services in low-income countries, especially from a societal perspective that captures household costs, remains limited.
2. Transitioning to WHO-recommended eight antenatal care visits in Uganda would increase total cost per woman by 57.8% to $157.9, with 63% of the incremental cost ($36.4) falling on households, equivalent to more than two-thirds of average monthly household income, and 37% ($21.4) on health facilities.
3. To ensure equitable scale-up, policy interventions should improve availability of medicines and diagnostics at public facilities, subsidize transport costs, and explore telemedicine or community-based models to reduce household financial burden.

## Background

In 2016, the World Health Organization (WHO) transitioned from a four-visit antenatal care (ANC) model to an eight-contact model ^1^, as part of broader global efforts to reduce maternal and neonatal mortality and achieve the United Nations Sustainable Development Goal 3 (SDG 3) of ensuring healthy lives and promoting well-being for all ^2^. This shift was based on increasing evidence that increased utilization of ANC has been associated with improved maternal and child health outcomes, including a significant reduction in neonatal mortality ^3^. However, for low-income countries, the variation between this global standard and local implementation capacity raises questions about affordability and feasibility. In these settings, the transition represents both a clinical and economic challenge for health systems and households.

While the Ministry of Health (MoH) has officially adopted the eight-visit model, achieving the previous four-visit standard remains a challenge. Only about 37% of pregnant women in Uganda attend their first ANC contact within the recommended first trimester, and one-third of women are unable to complete even four visits ^4^. With a maternal mortality ratio of 189 per 100,000 live births ^4^, it is important that pregnant women receive adequate monitoring throughout pregnancy. Yet doubling the required visit frequency requires a significant scale-up in service utilization for a population already facing barriers to access ^5^.

The capacity of the Ugandan health system to sustain this expansion depends on the country’s social and economic situation. Uganda ranks 157^th^ on the Human Development Index, with over 20% of the population living below the poverty line ^6^. In Uganda, out-of-pocket payments account for about 32% of total health spending ^7^, exceeding the 25% catastrophic expenditure threshold ^8^. With a national average monthly household income of around $105.0 ^9^, every additional visit to a health facility adds to this burden through transport fares and lost income from time away from work ^5^, pushing vulnerable households into deeper financial hardship. This situation is especially concerning given that the national health policy stipulates that health services at public facilities are free of charge at the point of use ^10^, yet patients frequently pay for services that are critical but unavailable at government facilities, such as ultrasound scans and laboratory tests.

Systemic financing challenges added to these individual-level barriers further complicate scaling efforts. Uganda currently allocates about 8.1% of its national budget to health ^11^, which is still below the 15% target established by the 2001 Abuja Declaration ^12^. Moreover, Uganda allocates about $45.2 per capita on health, below the WHO-recommended $86 required to deliver essential health services ^13,14^. The health sector depends heavily on donors, with external partners contributing nearly half of total national health expenditure ^13^, which creates uncertainty and complicates long-term planning for maternal and child health programs, especially following the scale-back in global health aid in 2025, whose implications for the health system are not yet known. In addition, the distribution of health funding in Uganda often prioritizes tertiary hospitals and administrative costs ^15,16^, leaving primary healthcare facilities, which handle the majority of ANC, struggling with shortages of essential medicines, diagnostic tools, and clinicians ^17^.

In addition to these systemic challenges, evidence on the incremental costs of scaling up ANC services in Uganda remains limited. As Hitimana et al. (2018) noted in their costing study of ANC in Rwanda, for countries considering adoption of the eight-visit model, it is necessary to understand the cost of the current practice, since without this baseline, policy decisions may not match the country’s available resources. Existing economic evaluations, such as a four-panel rapid screening test for infectious diseases, which was found to be cost-saving in the Ugandan context ^18^, have largely focused on the cost-effectiveness of clinical interventions, but do not address the financial implications of doubling service volume, a different type of intervention that increases service utilization rather than improving efficiency within existing visits. Moreover, there is limited evidence accounting for costs borne by patients, such as transport fares and lost productivity, despite studies identifying these as significant barriers to ANC attendance in Uganda ^5^.

Understanding these financial burdens is necessary to determine whether the eight-visit model is economically viable and equitable within an unpredictable financing landscape, a concern not only for Uganda but also other low-income countries facing similar constraints. Consequently, the objective of this study is to estimate the incremental costs to both the health system and patients of transitioning from the traditional four-visit ANC model to the WHO-recommended eight-visit model in Uganda.

## Methods

### Study setting and context

This study was conducted in Mbarara and Mitooma Districts in southwestern Uganda. The population is mostly peri-urban and rural, with agriculture being the primary economic activity. As is the case in the rest of the country, public facilities are organized in tiers according to roles and capacities ^19^. This system ranges from Health Center I (HC I), which consists of village health workers without a physical structure, to Health Center II (HC II), a nurse-led local dispensary. Health Center III (HC III) operates at the sub-county level, providing basic inpatient care under a clinical officer and midwives, and represents the lowest-level public facility offering antenatal and maternity services. Health Center IV (HC IV) functions at the county (sub-district) level, led by a medical officer, and delivers comprehensive services including emergency obstetric care. Above these are general hospitals at the district level, followed by regional referral hospitals and, at the highest level, the national referral hospital, which manages complex cases. Antenatal care services are available starting from HC III.

### Study design and overall approach

This study was nested within a randomized controlled trial (RCT) designed to evaluate a mobile health intervention aimed at promoting uptake of maternal health services ^20^. The analysis used a societal perspective, covering both health system costs and costs incurred at patients’ end to estimate the incremental cost of transitioning from current practice to the WHO-recommended eight-visit model, over a single pregnancy period. The current practice of ANC attendance in Uganda served as the comparator, with the WHO-recommended eight-visit model as the intervention of interest. The study used a micro-costing approach to identify, measure, and value all resources used in the provision of ANC services at selected government health facilities ^21^. This study was reported according to the Consolidated Health Economic Evaluation Reporting Standards (CHEERS) 2022 checklist ^22^.

### Facility selection

Six government health facilities were purposively selected across the two districts: two in Mitooma District and four in Mbarara District, comprising the largest HC III and HC IV in Mitooma District, and two HC IIIs and two HC IVs in Mbarara District. For the latter, facilities were selected from both Mbarara City and the district’s peri-urban areas, reflecting the diversity of the catchment population. The facility selection strategy was aimed at obtaining reliable cost estimates by capturing government facilities with sufficient patient volume to achieve economies of scale.

### Health facility costing

Data on health facility costs were collected using a data abstraction form adapted from Hitimana et al. (2018), through review of facility records and interviews with facility managers for the financial year of 2024/2025. Resources were categorized into four main input groups: personnel, equipment, supplies and consumables, and administrative overhead costs.

For personnel, information was collected on the number of staff involved in ANC activities, their cadres (e.g., medical officers, clinical officers, midwives, enrolled nurses), qualifications, and the level of effort allocated to ANC duties. Personnel salaries were obtained from the Uganda Public Service Commission salary structure ^23^, which provides standardized remuneration for public health workers.

Equipment costs included laboratory, furniture, computers, and other capital items used in ANC service delivery. Equipment was identified through interviews with facility staff, and unit prices were obtained from the MoH standard equipment price list ^24^. The annual cost of equipment was calculated by annuitizing the total value using a 3% discount rate, with the time allocated to ANC activities applied to derive the proportion attributable to ANC. Building and infrastructure costs were excluded as existing facilities were assumed to accommodate the additional visits without requiring new construction.

Supplies and consumables comprised drugs, laboratory reagents, vaccines (including tetanus toxoid), and other medical consumables used during ANC visits. Quantities were based on the standard MoH goal-oriented ANC protocol ^25^, which specifies the recommended tests and interventions for each visit. Prices for drugs and consumables were obtained from the National Medical Stores (NMS) and Joint Medical Stores (JMS) price lists issued to the respective facilities for the 2024/2025 financial year. These are internal procurement documents and are not publicly available. JMS is a large private supplier serving public facilities for items not available on the NMS list.

Administrative and overhead costs included utilities (electricity, water), cleaning services, telecommunications, stationery, and non-clinical staff salaries. These costs were extracted from the facility workplans and annual budgets. Overhead costs were allocated to ANC based on the proportion of ANC staff hours relative to total facility staff hours.

The total number of ANC visits, tests conducted, and cases of conditions tested were extracted from the registers of each facility for the year 2024/2025. Where data on test positivity rates were required to estimate costs conditional on positive cases, prevalence estimates were derived from facility registers. For HIV, cost estimates were obtained from published Ugandan literature due to the complexity of HIV-related services ^26^.

### Household costing

Costs incurred at patients’ end were collected using a healthcare utilization questionnaire administered to women after delivery as part of the parent RCT ^20^. The questionnaire captured information on all ANC visits during pregnancy, including the number of visits, the health providers seen, services received, costs incurred on transport and medical costs, as well as time spent to and from the facilities. Costs were disaggregated into three categories: transport costs (travel to and from the facility), ultrasound costs, and other medical costs (including medications and laboratory tests not available at government facilities). Sociodemographic characteristics were also collected, including age, distance from the health facility, monthly household income, and gestational age at enrollment.

The cost of transport was estimated based on the mean reported transport cost per visit, accounting for whether the woman was accompanied. Time costs were estimated based on travel and waiting time and valued using the human capital approach, with income used as a proxy for opportunity cost ^21^. Time costs were valued proportionally to monthly household income, and were similarly adjusted for accompaniment.

### Unit cost estimation

The unit cost per ANC visit was estimated for first visits and subsequent visits separately, as the content and intensity of services differ significantly. For facility costs, the total annual cost for each input category was divided by the total number of ANC visits to derive the average cost per visit. These costs were further disaggregated to estimate the cost of first visits and subsequent visits based on the respective resource use specified in the MoH goal-oriented ANC protocol ^25^. For patient costs, the average cost per first visit and subsequent visit was calculated from our patient survey data. Total patient costs of ANC per woman were estimated by initially calculating the cost of the first visit and multiplying the per-visit costs by the average number of visits attended less one.

### Projection of the WHO-recommended eight-visit model costs

To estimate the incremental cost of transitioning to the WHO-recommended eight-visit model, the cost of the current four-visit model was scaled up to eight visits. For facility costs, the additional visits were assumed to follow the same resource intensity as subsequent visits (rather than first visits), consistent with the MoH protocol in which only the first visit includes the full battery of laboratory tests and intensive assessments. For patient costs, transport and medical costs were multiplied by the additional number of visits, while ultrasound costs were projected based on the average number of scans per woman in the current practice and the assumption that ultrasound utilization patterns would remain consistent.

### Cost analysis and sensitivity analysis

All costs were analyzed in Ugandan shillings and converted to United States dollars using the Bank of Uganda exchange rate at the close of the 2024/2025 financial year (1 USD = 3599.4 UGX, June 2025). Analysis was conducted by facility level, given the variation in cost structures, especially in personnel composition and patient volumes across facilities. Patient costs were similarly disaggregated by facility level to reflect these differences. Missing data for time to and from the facility, number of ultrasound scans, and ultrasound costs were addressed by replacing missing values with facility-level means.

We performed a one-way sensitivity analysis to assess the robustness of the incremental cost estimates to key assumptions ^21^. For patient costs, parameters varied including transport cost per visit, ultrasound scans per patient, medical costs per visit, time cost per visit, and first visit intensity, with ranges derived from the 95% confidence intervals obtained from study participant data. For facility costs, parameters that were varied included equipment lifetime based on individual equipment’s estimated lifetime, discount rate (0-5%), personnel level of effort (+/-20%), consumables cost per visit (+/- 20%), and first visit intensity (+/- 20%) ^27^. Each parameter was varied independently while holding all others at base case values. Results were presented using tornado diagrams to illustrate the relative impact of each parameter on incremental facility and patient costs.

### Ethical approval

Ethical approval for the study was obtained from Mbarara University of Science and Technology Research Ethics Committee under reference number MUST-2025-587. Written informed consent was obtained from all participants before enrollment in the parent RCT. This consent included the use of data for economic analyses.

## Results

### Facility and participant characteristics

Data collection was conducted in six government health facilities, including three HC IVs (Bwizibwera, Mitooma, and Mbarara Municipal Council) and three HC IIIs (Rubindi, Bitereko, and Kakoba). Table 1 presents the characteristics of the study facilities and patients’ sample. A total of 9,784 women received ANC during the one-year study period, contributing 42,075 visits with an average of 4.3 visits per woman. The total annual facility cost for ANC services to run these six facilities was $427,767.

**Table 1:**
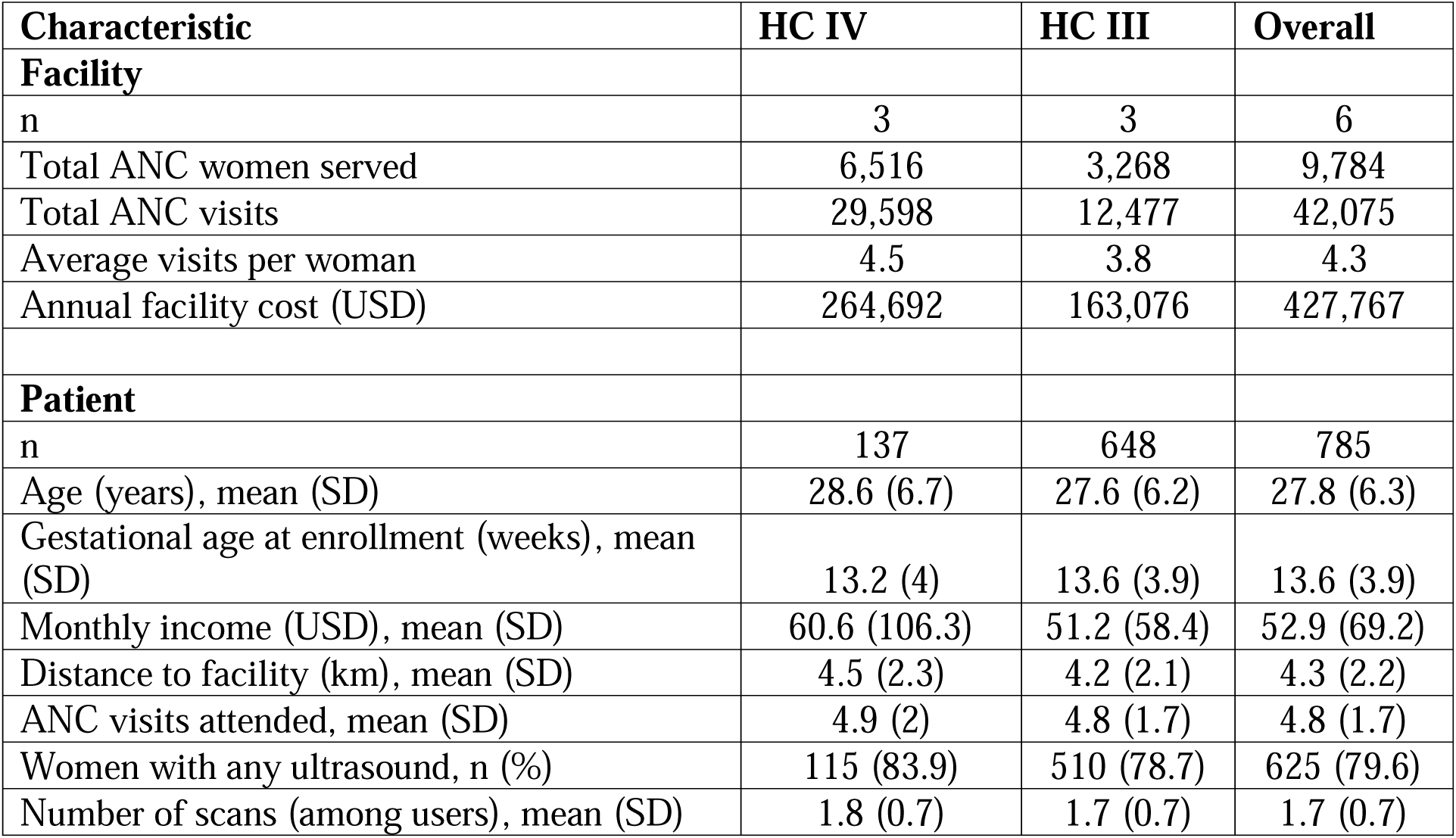
Characteristics of study facilities and patient sample, Mbarara and Mitooma Districts, Southwestern Uganda, 2024/2025 (in 2025 USD)

| Characteristic | HC IV | HC III | Overall |
| --- | --- | --- | --- |
| **Facility** |  |  |  |
| n | 3 | 3 | 6 |
| Total ANC women served | 6,516 | 3,268 | 9,784 |
| Total ANC visits | 29,598 | 12,477 | 42,075 |
| Average visits per woman | 4.5 | 3.8 | 4.3 |
| Annual facility cost (USD) | 264,692 | 163,076 | 427,767 |
| **Patient** |  |  |  |
| n | 137 | 648 | 785 |
| Age (years), mean (SD) | 28.6 (6.7) | 27.6 (6.2) | 27.8 (6.3) |
| Gestational age at enrollment (weeks), mean (SD) | 13.2 (4) | 13.6 (3.9) | 13.6 (3.9) |
| Monthly income (USD), mean (SD) | 60.6 (106.3) | 51.2 (58.4) | 52.9 (69.2) |
| Distance to facility (km), mean (SD) | 4.5 (2.3) | 4.2 (2.1) | 4.3 (2.2) |
| ANC visits attended, mean (SD) | 4.9 (2) | 4.8 (1.7) | 4.8 (1.7) |
| Women with any ultrasound, n (%) | 115 (83.9) | 510 (78.7) | 625 (79.6) |
| Number of scans (among users), mean (SD) | 1.8 (0.7) | 1.7 (0.7) | 1.7 (0.7) |

The patients’ sample comprised 785 women, with a higher proportion attending HC IIIs, reflecting the larger catchment populations served by lower-tier facilities in this predominantly rural and peri-urban setting. The average visits per woman was slightly higher in the patient sample (4.8) than in facility records (4.3), as facility data includes all women attending ANC beyond those enrolled in the study. Monthly household income averaged $52.9 (SD 69.2), and 80% of women reported at least one ultrasound scan, with an average of 1.7 scans among users.

### Current cost of ANC

Table 2 presents the current cost of ANC under the current practice, disaggregated by facility tier and by visit type. The total cost per woman for the full course of ANC averaged $100.1 across all facilities. Facility costs accounted for $43.7 (43.7%) and costs incurred at patients’ end accounted for $56.4 (56.3%).

**Table 2:**
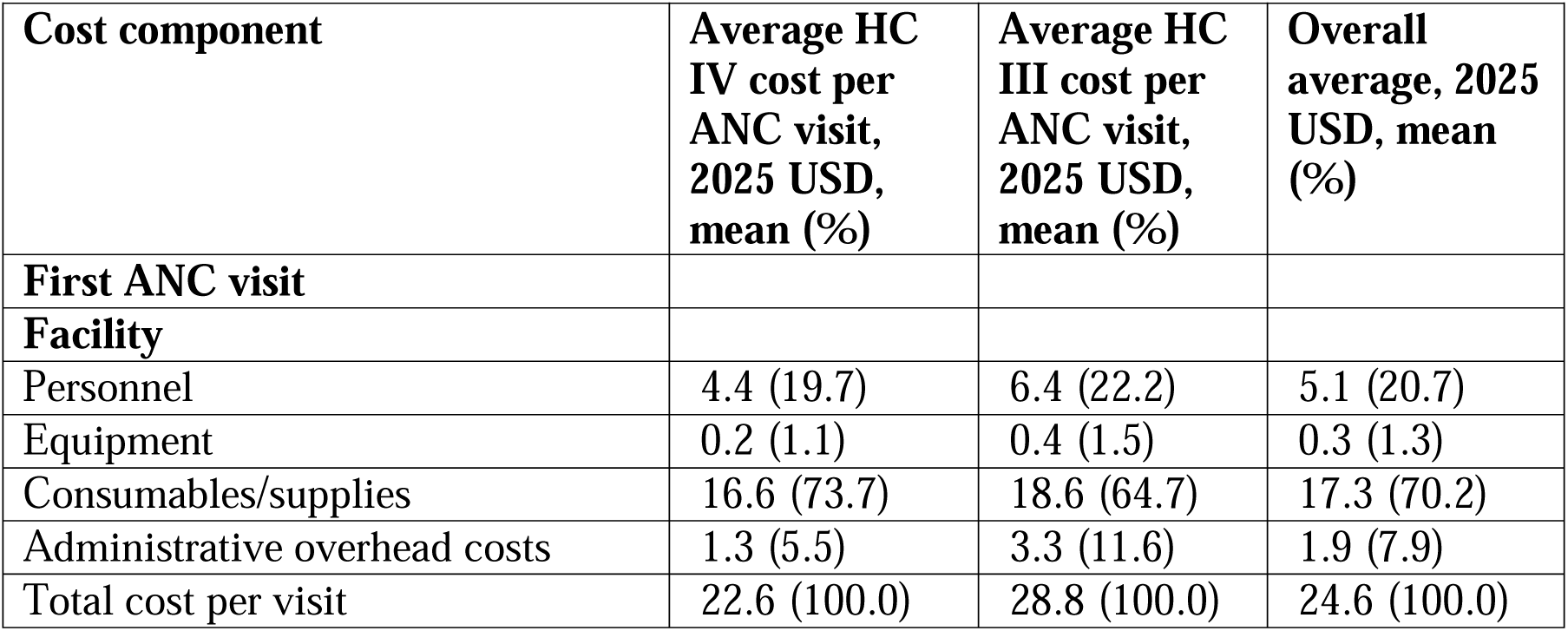
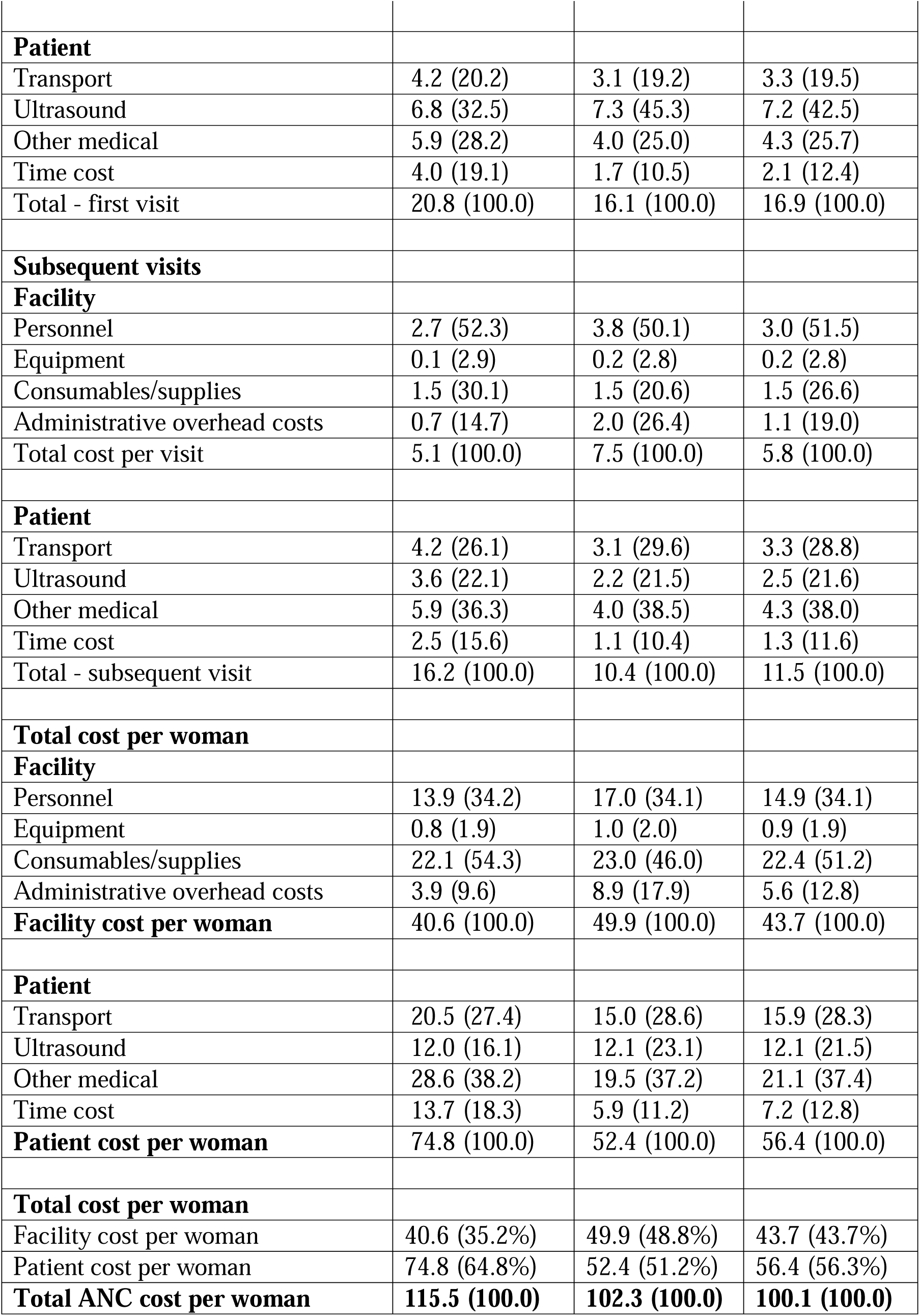
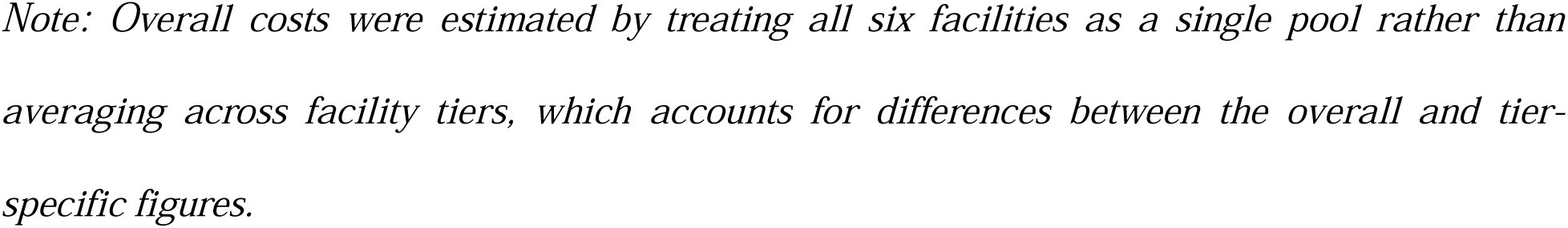
Current cost of ANC per woman, Mbarara and Mitooma Districts, Southwestern Uganda, 2024/2025 (in 2025 USD)

| Cost component | Average HC IV cost per ANC visit, 2025 USD, mean (%) | Average HC III cost per ANC visit, 2025 USD, mean (%) | Overall average, 2025 USD, mean (%) |
| --- | --- | --- | --- |
| **First ANC visit** |  |  |  |
| **Facility** |  |  |  |
| Personnel | 4.4 (19.7) | 6.4 (22.2) | 5.1 (20.7) |
| Equipment | 0.2 (1.1) | 0.4 (1.5) | 0.3 (1.3) |
| Consumables/supplies | 16.6 (73.7) | 18.6 (64.7) | 17.3 (70.2) |
| Administrative overhead costs | 1.3 (5.5) | 3.3 (11.6) | 1.9 (7.9) |
| Total cost per visit | 22.6 (100.0) | 28.8 (100.0) | 24.6 (100.0) |
| **Patient** |  |  |  |
| Transport | 4.2 (20.2) | 3.1 (19.2) | 3.3 (19.5) |
| Ultrasound | 6.8 (32.5) | 7.3 (45.3) | 7.2 (42.5) |
| Other medical | 5.9 (28.2) | 4.0 (25.0) | 4.3 (25.7) |
| Time cost | 4.0 (19.1) | 1.7 (10.5) | 2.1 (12.4) |
| Total - first visit | 20.8 (100.0) | 16.1 (100.0) | 16.9 (100.0) |
| **Subsequent visits** |  |  |  |
| **Facility** |  |  |  |
| Personnel | 2.7 (52.3) | 3.8 (50.1) | 3.0 (51.5) |
| Equipment | 0.1 (2.9) | 0.2 (2.8) | 0.2 (2.8) |
| Consumables/supplies | 1.5 (30.1) | 1.5 (20.6) | 1.5 (26.6) |
| Administrative overhead costs | 0.7 (14.7) | 2.0 (26.4) | 1.1 (19.0) |
| Total cost per visit | 5.1 (100.0) | 7.5 (100.0) | 5.8 (100.0) |
| **Patient** |  |  |  |
| Transport | 4.2 (26.1) | 3.1 (29.6) | 3.3 (28.8) |
| Ultrasound | 3.6 (22.1) | 2.2 (21.5) | 2.5 (21.6) |
| Other medical | 5.9 (36.3) | 4.0 (38.5) | 4.3 (38.0) |
| Time cost | 2.5 (15.6) | 1.1 (10.4) | 1.3 (11.6) |
| Total - subsequent visit | 16.2 (100.0) | 10.4 (100.0) | 11.5 (100.0) |
| **Total cost per woman** |  |  |  |
| **Facility** |  |  |  |
| Personnel | 13.9 (34.2) | 17.0 (34.1) | 14.9 (34.1) |
| Equipment | 0.8 (1.9) | 1.0 (2.0) | 0.9 (1.9) |
| Consumables/supplies | 22.1 (54.3) | 23.0 (46.0) | 22.4 (51.2) |
| Administrative overhead costs | 3.9 (9.6) | 8.9 (17.9) | 5.6 (12.8) |
| **Facility cost per woman** | 40.6 (100.0) | 49.9 (100.0) | 43.7 (100.0) |
| **Patient** |  |  |  |
| Transport | 20.5 (27.4) | 15.0 (28.6) | 15.9 (28.3) |
| Ultrasound | 12.0 (16.1) | 12.1 (23.1) | 12.1 (21.5) |
| Other medical | 28.6 (38.2) | 19.5 (37.2) | 21.1 (37.4) |
| Time cost | 13.7 (18.3) | 5.9 (11.2) | 7.2 (12.8) |
| **Patient cost per woman** | 74.8 (100.0) | 52.4 (100.0) | 56.4 (100.0) |
| **Total cost per woman** |  |  |  |
| Facility cost per woman | 40.6 (35.2%) | 49.9 (48.8%) | 43.7 (43.7%) |
| Patient cost per woman | 74.8 (64.8%) | 52.4 (51.2%) | 56.4 (56.3%) |
| **Total ANC cost per woman** | **115.5 (100.0)** | **102.3 (100.0)** | **100.1 (100.0)** |

*Note: Overall costs were estimated by treating all six facilities as a single pool rather than averaging across facility tiers, which accounts for differences between the overall and tier-specific figures.*

The first visit was significantly more costly than subsequent visits, largely due to consumables and supplies at the facility level and ultrasound utilization at the patient level. Consumables and supplies accounted for $17.3 (70.2%) of first visit facility costs compared to $1.5 (26.6%) of subsequent visit facility costs. Ultrasound costs declined from $7.2 at the first visit to $2.5 at each subsequent visit.

Costs differed by facility tier. Facility costs per woman were higher at HC IIIs ($49.9) than at HC IVs ($40.6), driven by higher administrative overhead costs (17.9% vs 9.6% of facility costs), reflecting the lower patient volumes over which fixed overhead costs were distributed. In contrast, patient costs per woman were higher at HC IVs ($74.8) than at HC IIIs ($52.4), driven primarily by higher medical costs ($28.6 vs $19.5) and higher time costs ($13.7 vs $5.9). Across all facilities, consumables and supplies were the largest component of facility costs, accounting for 46.0–54.3% of total facility costs per woman. Personnel costs were the second largest category, representing 34.1% of facility costs. Equipment cost contributed a minor share (2.0%) across both facility tiers.

For patient costs, other medical costs comprising medications and tests not available at government facilities constituted the largest category, accounting for 37.2–38.2% of total patient costs per woman. Ultrasound costs represented 16.1–23.1% of patient costs, while transport accounted for 27.4–28.6%.

### Projected cost of the WHO-recommended eight-visit model

Table 3 presents the projected cost of scaling up to the WHO-recommended eight-visit model, under which the total cost per woman across all facilities is projected to increase by 57.8% to $157.9. The incremental cost per woman is estimated at $57.8, comprising $21.4 (37.0%) in additional facility costs and $36.4 (63.0%) in additional costs incurred at patients’ end.

**Table 3:** Projected cost of WHO-recommended 8 ANC visits by facility tier (USD)

| **Cost component** | **Average HC IV cost per ANC visit, 2025 USD, mean (%)** | **Average HC III cost per ANC visit, 2025 USD, mean (%)** | **Overall average, 2025 USD, mean (%)** |
| --- | --- | --- | --- |
| **Facility** |  |  |  |
| Personnel | 23.1 (39.7) | 32.7 (40.3) | 25.9 (39.8) |
| Equipment | 1.3 (2.2) | 1.9 (2.3) | 1.5 (2.2) |
| Consumables/supplies | 27.4 (47.0) | 29.4 (36.2) | 28.1 (43.1) |
| Administrative overhead costs | 6.5 (11.2) | 17.2 (21.2) | 9.7 (14.8) |
| **Facility cost per woman** | **58.2 (100.0)** | **81.3 (100.0)** | **65.1 (100.0)** |
| **Patient** |  |  |  |
| Transport | 33.7 (27.8) | 24.8 (28.5) | 26.3 (28.4) |
| Ultrasound | 19.0 (15.7) | 20.6 (23.7) | 20.3 (21.8) |
| Other medical | 47.0 (38.7) | 32.2 (37.1) | 34.8 (37.5) |
| Time cost | 21.6 (17.8) | 9.3 (10.7) | 11.4 (12.3) |
| **Patient cost per woman** | **121.3 (100.0)** | **86.8 (100.0)** | **92.8 (100.0)** |
| **Total cost per woman** |  |  |  |
| Facility cost per woman | 58.2 (32.4%) | 81.3 (48.4%) | 65.1 (41.2%) |
| Patient cost per woman | 121.3 (67.6%) | 86.8 (51.6%) | 92.8 (58.8%) |
| **Total ANC cost per woman** | **179.5 (100.0)** | **168.1 (100.0)** | **157.9 (100.0)** |

*Note: Overall costs were estimated by treating all six facilities as a single pool rather than averaging across facility tiers, which accounts for differences between the overall and tier-specific figures.*

As shown in Figure 1, the total cost per woman at HC IVs would increase by 55.4% (facility 43.4%, patient 61.2%), with 72.5% of the incremental cost falling on patients. At HC IIIs, the total cost per woman would increase by 64.3% (facility 62.9%, patient 65.7%), with the incremental cost more evenly split between facilities (47.7%) and patients (52.3%).

**Figure 1:**
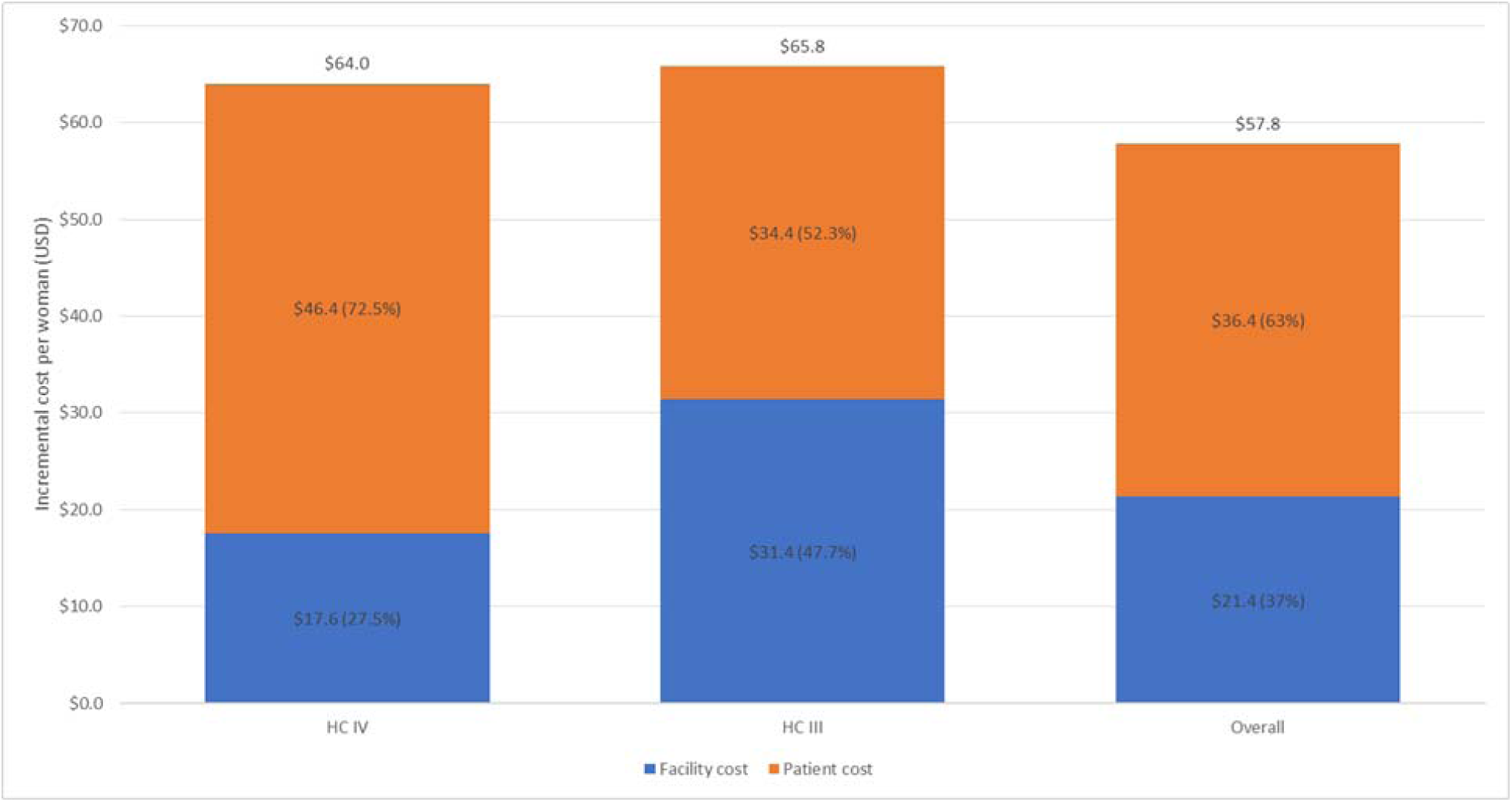
Incremental cost per woman of transitioning from current practice to WHO-recommended 8 ANC visits, by facility tier and cost bearer (2025 USD)

As presented in Figure 2, one-way sensitivity analysis showed that the incremental cost per woman was stable across tested parameters. For patient costs, medical costs had the largest impact ($34.9–$38.0), while transport, ultrasound, time costs, and first visit intensity had smaller impacts. For facility costs, personnel level of effort had the largest impact ($19.2–$23.6), followed by first visit intensity and consumables costs. Equipment lifetime and discount rate had minimal impact.

**Figure 2:**
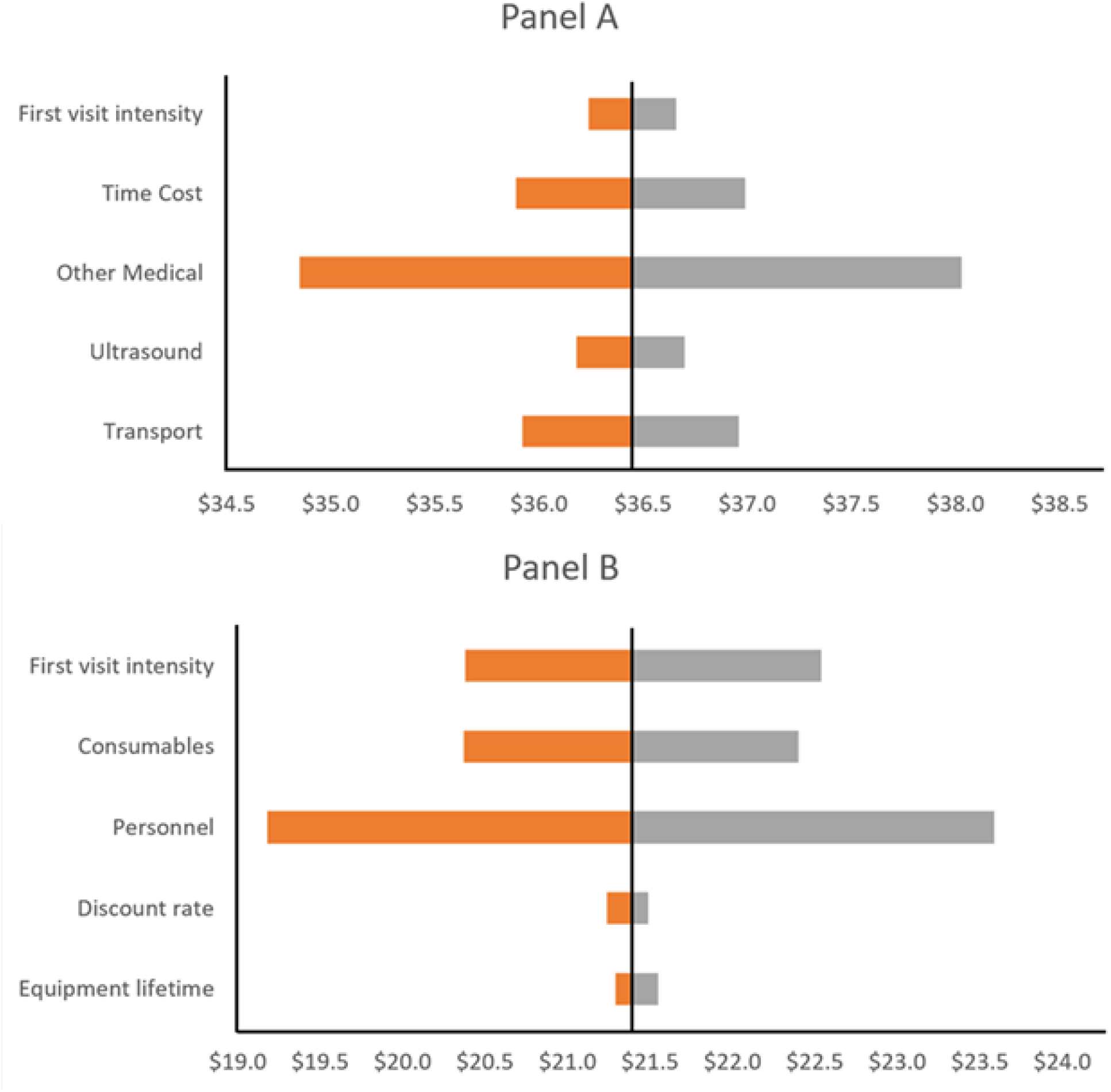
One-way sensitivity analysis for incremental patient and facility costs. Tornado diagrams show incremental patient costs (Panel A) and incremental facility costs (Panel B). The vertical line represents the base case incremental cost per woman ($36.4 for patient costs; $21.4 for facility costs). Bars show the range of incremental cost per woman when each parameter is varied between its lower and upper bounds. The category “Other Medical” includes medicines, diagnostics, and laboratory services not available at public facilities

## Discussion

Transitioning to eight visits would increase total cost per woman by 57.8% to $157.9, with $36.4 (63.0%) of the incremental cost falling on households, equivalent to more than two-thirds of average monthly household income and above the 25% catastrophic expenditure threshold ^8^. This burden is especially concerning among women in this setting, whose average monthly household income of $52.9 is well below the national average of $105.0, and in a country where 32% of total health spending comes directly from households ^7^. Despite this cost burden, the eight-visit model was designed to strengthen monitoring of hypertensive disorders and other asymptomatic complications, and evidence has shown more frequent ANC to be associated with a 34% reduction in the risk of neonatal mortality, as well as a positive pregnancy and delivery experience ^1,3^. In Uganda, where maternal mortality remains high at 189 per 100,000 live births ^4^, this incremental cost may be worthwhile given clinical benefits, provided that the financial barriers identified in this study are addressed to ensure equitable access.

Facility costs also differed by tier. HC IIIs had higher facility costs per woman ($49.9) than HC IVs ($40.6), a difference driven primarily by administrative overhead costs (17.9% vs 9.6%), reflecting lower patient volumes over which fixed costs are distributed. The incremental facility cost of scaling up to eight visits was also projected to be higher at HC IIIs (62.9%) than at HC IVs (43.4%), suggesting that lower-level facilities may struggle to sustain increased service volume. The high costs incurred at patients’ end observed across both facility tiers, largely due to expenditures on medicines and diagnostics that should be available free of charge at government facilities, reflect a wider problem in Uganda’s health financing landscape, where supply chain challenges and inadequate government allocations leave primary healthcare facilities under-resourced ^15,17^. Improving the availability of essential medicines and diagnostics at public facilities would therefore reduce both household burden and the equity gap between facility tiers. Since HC IIIs are more accessible to the majority of women in this setting, strengthening capacity at this level through task shifting and improved scheduling may be a more equitable and cost-efficient strategy than subsidizing the higher patient costs associated with attendance at HC IVs.

The challenge of scaling up ANC services in Uganda must also be seen in the context of the country’s health financing constraints. The health sector relies heavily on donor funds, which creates financing unpredictability, a concern heightened by the scale-back in global health aid in 2025 ^13^. This funding reality means that doubling the number of ANC visits competes directly with other national health priorities in an already overstretched system struggling with inadequate staffing, essential medicine stockouts, and limited diagnostic equipment even for the current four visits, let alone eight ^15,17^. With one-third of women still unable to complete even four visits ^4^, the country has yet to achieve universal coverage of the previous model despite years of policy commitment. Expanding to eight contacts without appropriate investment in health system capacity could produce a policy that is adopted but not implemented.

Our results are broadly comparable to a costing study of ANC in Rwanda, which found a societal cost of $44 per woman for four visits, with facility costs of $42.6 ^28^, although with notable differences in cost composition. While facility costs were similar, patient costs are higher in our study at $56.4 per woman, representing 57% of the total cost compared to $1.4 (3%) per woman in Rwanda. The difference can be explained by the Rwanda study’s inclusion of only transport and time costs, which may have underestimated the patients’ financial burden. Our study has the strength of additionally capturing ultrasound and other medical costs, which together represented 59% of patient costs. Future costing studies in similar settings need to adopt a comprehensive societal perspective that captures all out-of-pocket expenditures, as facility-perspective analyses alone may underestimate the true financial burden on patients.

## Limitations

The study was conducted in two districts in southwestern Uganda, and so the findings may not be generalizable to some other parts of the country with different cost structures. Additionally, the study was nested within an RCT evaluating a mobile health intervention; thus, some of the eligibility criteria (i.e., having a supporting caregiver) may have made the study population slightly different from the general population.

The projection of eight-visit costs assumes uniform resource intensity across subsequent visits. Although the MoH goal-oriented ANC protocol specifies differentiated goals across visits, individual visit-level cost data were not routinely captured in facility records, as many tests and interventions are symptom-driven rather than visit-specific, making it difficult to attribute costs to individual visit numbers. Nevertheless, later visits, especially in the third trimester, may involve higher-intensity care, meaning our estimates are likely conservative.

Patient cost data relied on self-reported recall over up to nine months, which may introduce slight recall bias. However, women were asked about costs they repeatedly incurred on the same journey to a familiar facility, as well as ultrasound and other medical costs they likely kept records of, suggesting recall was likely adequate. Finally, the study excluded private and faith-based facilities, and findings might not apply to non-public providers.

## Conclusion

Transitioning from four to eight ANC visits in Uganda would require an additional $57.8 per woman, with 63.0% of this burden falling on households, equivalent to more than two-thirds of average monthly household income of $52.9. With only 8.1% of the national budget allocated to health, the government faces difficult choices about where to direct limited health funds.

An important step is to improve the availability of essential medicines and diagnostics at public facilities, given that these were the largest driver of patient costs and the most sensitive parameter in the analysis. Targeted transport subsidies or vouchers for women living further from facilities would address transport costs, which accounted for 27.4–28.6% of patient costs. Efficiency measures at HC IIIs, including task shifting and improved scheduling, could contribute to reduced costs of scale-up. Finally, telemedicine or community-based contact models could reduce transport and time costs associated with physical attendance for some of the additional contacts where clinically appropriate.

While the clinical benefits of increased ANC (including positive pregnancy and delivery experience, pronounced decrease in neonatal mortality) are well documented, the feasibility of adopting the eight-visit model in Uganda depends on whether the health system can absorb the additional facility costs and whether households can bear the associated financial burden.

## Data availability

The datasets used and analysed during the current study are available from the corresponding author on reasonable request.

## Data Availability

All data produced in the present study are available upon reasonable request to the authors

## Notes

### Competing Interest Statement

The authors have declared no competing interest.

### Clinical Trial

NCT05940831

### Author Declarations

Mbarara University of Science and Technology Research Ethics Committee. Approval granted under reference number MUST-2025-587

### Funding

- National Institutes of Health [NIH]; Eunice Kennedy Shriver National Institute of Child Health & Human Development: R01HD111692

